# Variable association of atypical femur fracture and osteonecrosis of jaw with bisphosphonates and denosumab use: Drug-safety surveillance study

**DOI:** 10.1101/2020.06.15.20132290

**Authors:** Tigran Makunts, Lara S. Anwar, Ruben Abagyan

## Abstract

In the United States, there are over ten million adults diagnosed with osteoporosis and many more are at risk of developing the condition. Osteoporosis affects both males and females, mostly post-menopausal. Bisphosphonates and denosumab have been widely used globally to treat the condition. The use of bisphosphonates and denosumab had been associated with rare adverse effects including osteonecrosis of the jaw, ONJ, and atypical femur fracture, AFF. However, it remained unclear whether those side effects were class-wide or drug-specific. By analyzing over 230,000 osteoporosis patient reports from the FDA adverse event reporting system, FAERS, we confirmed the association of bisphosphonates and denosumab use with AFF and ONJ. Additionally, comparing each of the four frequently used bisphosphonates with denosumab-treated patients used as a control, we identified: (i) varying significance of association with ONJ and AFF for alendronate, risedronate, ibandronate and zoledronic acid, (ii) over two fold increase in risk of both side effects in alendronate patients, particularly in females, (iii) over a six fold increase in AFF risk in both males and females taking risedronate, and (iv) lower risk of both AFF and ONJ, for zoledronic acid patients compared to denosumab.

**Key points:**

1. We performed a disproportionality analysis of over 230,000 post-marketing reports of patients treated for osteoporosis to measure the risk of developing atypical femur fracture (AFF) and osteonecrosis of the jaw (ONJ).
2. Alendronate, ibandronate, risedronate, zoledronic acid, and denosumab were all significantly associated with AFF and ONJ when compared to teriparatide.
3. When compared to denosumab, patients taking alendronate, ibandronate, risedronate, or zoledronic acid had a variable risk of ONJ and AFF, which correlated with the frequency of drug administration. The trend in variable risk was observed in both females and males.

## Introduction

The National Osteoporosis Foundation reports that 10.2 million adults age fifty and older had osteoporosis in the U.S in 2013, and 43.4 million adults were at risk of developing the disease, giving a total of about 54 million U.S. adults with osteoporosis or low bone mass[1]. It was projected that this total number will grow to 64.4 million by 2020 and to 71.2 million by 2030[1]. Moreover, the study showed that more women than men had osteoporosis (8.2 million vs. 2.0 million) and low bone mass (27.3 million vs. 16.1 million)[1]. Osteoporosis causes a reduction of bone mass and strength due to multiple pathogenic mechanisms, leading to increased bone fragility and susceptibility to breakage[2, 3].

Three main types of drugs are recommended for the treatment of osteoporosis, including bisphosphonates (alendronate, risedronate, ibandronate, and zoledronic acid), denosumab and teriparatide[4]. Bisphosphonates are small molecules that bind to hydroxyapatite in bone, leading to the inhibition of osteoclastic bone resorption[5, 6], denosumab is a fully humanized monoclonal antibody to RANKL[7], and teriparatide is a synthetic version of the human parathyroid hormone which works by activating osteoblasts[8, 9].

Common adverse effects of oral bisphosphonates (alendronate, risedronate, ibandronate) include, but not limited to, upper gastrointestinal side effects (acid reflux, esophagitis, ulcers) due to local effects on esophagus and/or gastric mucosa[10-12]. Intravenous (IV) bisphosphonates (ibandronate, zoledronic acid) are associated with injection-related reactions, such as, flu-like symptoms (fever, myalgias, arthralgias)[12, 13]. Musculoskeletal pain is common to both oral and IV formulations[10-13].

Denosumab, administered as a subcutaneous injection, is associated with fatigue/asthenia, hypophosphatemia, nausea, and back pain[14]. Common side effects of teriparatide include injection-site pain, nausea, headaches, leg cramps, and dizziness[15].

While these adverse effects are expected and can be managed, there are two counterintuitive and troubling adverse effects, unique to bisphosphonates and denosumab, that directly damage the bone instead of preventing bone loss: 1) osteonecrosis of the jaw (ONJ), and 2) atypical femur fracture (AFF)[16].

AFF is referred to as “atypical” due to the location of the fracture. AFF occurs in the subtrochanteric or diaphyseal region of the femur with simple transverse or oblique pattern[16]. The pathophysiology of AFF is not well understood. However, it is suggested that since bisphosphonates inhibit the activity of osteoclasts, they cause suppression of bone turnover rate, resulting in the effects that contribute to AFF, such as accumulation of microdamage in the bone[16]. Similarly, denosumab provokes the disruption of the targeted remodeling process, which is needed to replace microcracks with new bone tissue, leading to accumulation of bone microdamage[17]. A study done by Black and colleagues reviewed 284 records of hip or femur fractures associated with bisphosphonates use among 14,195 women in these trials[18]. The study showed no significant increase in the risk of AFF for alendronate use (relative risk (RR) 1.03, 95% confidence interval (CI) [0.06-16.46]) and for zoledronic acid use (RR 1.50, 95% CI [0.25-9.00]) compared to placebo[18]. However, many case reports and small-scale studies have shown the association of bisphosphonate use with AFF for alendronate, ibandronate, risedronate, zoledronic acid, and denosumab[19-27].

ONJ is a severe disease that affects the jaw, and it is defined as exposed, necrotic bone in the maxillary and mandibular regions[28]. There are multiple factors that play a role in the pathophysiology of ONJ, putting the oral cavity at a higher risk[29]. Oral structures can be subjected to various types of stresses such as mastication, dental procedures, periodontal disease, caries, poor oral hygiene and effects of chemotherapy[29]. Combination of these factors can lead to bone exposure and demand higher rates of bone remodeling[29]. It is advised for all patients to undergo dental examination prior to initiating treatment with bisphosphonates or denosumab, as well as, maintaining good oral hygiene throughout the treatment[30].

A literature review of eleven publications reporting 26 cases of ONJ was performed by Pazianas and colleagues to clarify the association between bisphosphonate use and the development of such adverse event[31]. The study showed that the relative prevalence of ONJ was low and more often associated with a dental procedure; however, the authors did not draw any conclusion related to the differences between the drugs due to the scarcity of the reports[31]. There have been a few case reports and small-scale studies on the ONJ ADR in patients taking alendronate, ibandronate, risedronate, zoledronic acid, and denosumab[32-37].

To further quantify the drug-specific associations those treatments with AFF and ONJ and expand on current evidence we analyzed over twelve million post-marketing adverse drug reaction reports. Initially, we illustrated the association of ONJ and AFF with bisphosphonates and denosumab in relation to teriparatide. Teriparatide was chosen due to its unique mechanism of action and AE profile [38-40].

Additionally, we evaluated the reporting odds ratios (RORs) of AFF and ONJ in patients with osteoporosis for each of four bisphosphonates using denosumab as a control and evaluated the co-occurrence of these AEs. The magnitude of the effects, and their 95% confidence intervals (CIs) were also evaluated for each bisphosphonate in both male and female patients to quantify additional risk factors.

## Methods

FAERS/AERS contains reports of medication-related adverse effects submitted to the FDA voluntarily by patients, healthcare providers, and legal representatives through MedWatch[41], the FDA Safety Information and Adverse Event Reporting System. FAERS reports were utilized to conduct a retrospective analysis on patients with osteoporosis, taking either bisphosphonates, denosumab, or teriparatide to compare the frequency of rare adverse events, AFF and ONJ, associated with the use of these drugs.

Data sets are available to the public online at: http://www.fda.gov/Drugs/GuidanceComplianceRegulatoryInformation/Surveillance/AdverseDrugEffects/ucm082193.htm.

### Normalizing and combining the data

Each quarterly report set was then downloaded in a (.TXT) format and modified to produce a standardized table field structure. Some data sets contained missing columns, and blank columns were added with no values to homogenize the reports[42, 43]. The final data set contained over twelve million reports. Reports were submitted mostly from the United States, however, many reports were submitted from around the world with their country-specific formats. International brand and generic names were translated into a single generic form using online drug databases[42, 43].

### Choosing the cohorts

The FAERS system was queried based on the adverse events reported between January of 2004 and March of 2019. A total of 12,004,552 FAERS adverse reaction reports were collected. Reports of patients with osteoporosis were selected into the osteoporosis cohort if the “indication” field in the FAERS reports had the term osteoporosis exclusively. Approximately 0.04% of FAERS reports are duplicates. These are follow-up reports with the same case numbers. In our case selection, the duplicate reports were excluded. Additionally, reports by lawyers were also excluded due to potential bias.

Guidelines and position statements from the following organizations and societies were used to identify the first-line recommended therapeutics for osteoporosis prevention and treatment: The American Association of Clinical Endocrinologists[44], the Endocrine Society[45], the American Academy of Family Physicians[46], the National Osteoporosis Foundation[47], and the North American Menopause Society[48]. Alendronate, ibandronate, risedronate, zoledronic acid, and denosumab were all recommended as a first-line line therapeutic option.

From 232,512 osteoporosis reports, a total of 133,089 osteoporosis monotherapy reports were selected. Out of the latter osteoporosis group, reports where denosumab was used for the treatment of osteoporosis, excluding bisphosphonates (alendronate, ibandronate, risedronate, zoledronic acid) and other drugs associated with ONJ such as m-TOR inhibitors (sirolimus, everolimus, temsirolimus), and antiangiogenic drugs (bevacizumab, sunitinib, sorafenib, pazopanib, axitinib), were selected into the denosumab cohort (n=18,336). Reports where bisphosphonates were used, excluding denosumab, m-TOR inhibitors, and antiangiogenic drugs, were selected into the bisphosphonates cohort (n=36,527). Out of the osteoporosis reports, reports where teriparatide were used for the treatment of osteoporosis, excluding denosumab, bisphosphonates, m-TOR inhibitors, and antiangiogenic drugs, were selected into the teriparatide cohort (n=66,173). The bisphosphonate cohort was further split into alendronate (n=14,682), ibandronate (n=6,065), risedronate (n=2,309), and zoledronic acid (n=13,471) sub-cohorts (Fig 1). Demographic analysis was also performed (Tables 1 and 2).

**Table 1.**
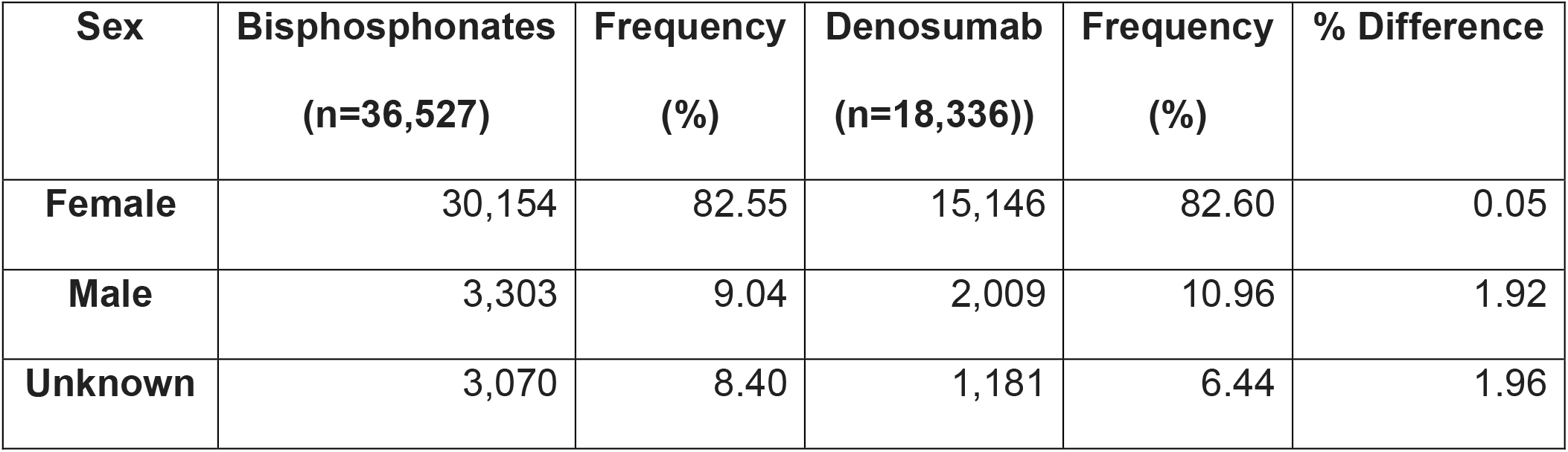
Patient demographics in bisphosphonates and denosumab cohorts.

| **Sex** | **Bisphosphonates**<br>**(n=36,527)** | **Frequency**<br>**(%)** | **Denosumab**<br>**(n=18,336))** | **Frequency**<br>**(%)** | **% Difference** |
| --- | --- | --- | --- | --- | --- |
| **Female** | 30,154 | 82.55 | 15,146 | 82.60 | 0.05 |
| **Male** | 3,303 | 9.04 | 2,009 | 10.96 | 1.92 |
| **Unknown** | 3,070 | 8.40 | 1,181 | 6.44 | 1.96 |

**Table 2.** Age difference in patients with osteoporosis taking bisphosphonates vs. denosumab.

|  | **Bisphosphonates** | **Denosumab** |
| --- | --- | --- |
| **Mean Age, Years (SD)** | 69.10 (12.22) | 73.01 (11.52) |
| **Median Age, Years** | 69.40 | 67.60 |
| **Unknown (%)** | 32.60 | 25.98 |

**Fig 1.**
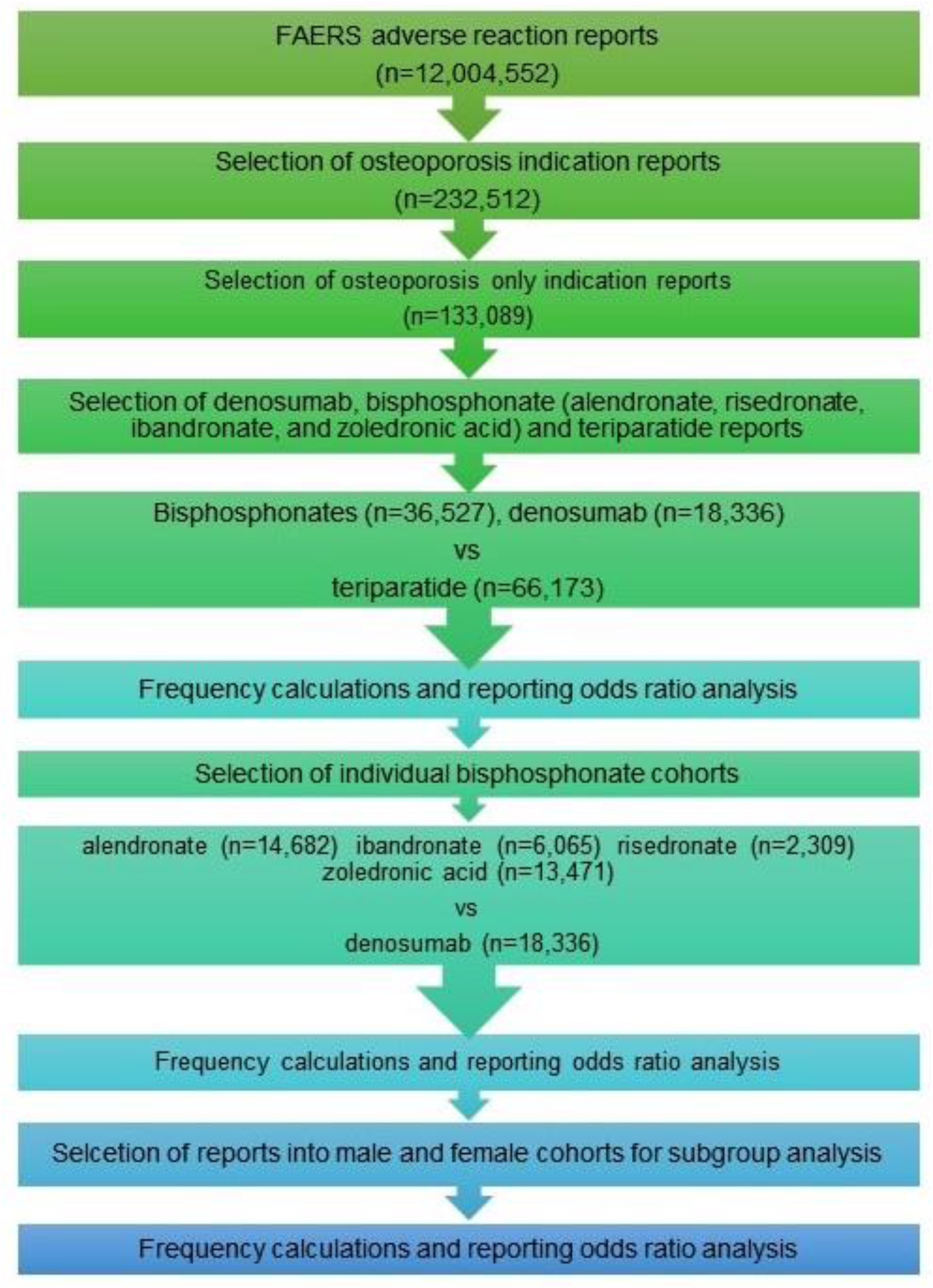
Cohort selection. Cohort selection for adverse event rate comparison between bisphosphonates, denosumab and teriparatide as a class, and between individual bisphosphonates and denosumab. Abbreviations: FAERS – FDA Adverse Event Reporting System,

### Statistical analysis

#### Descriptive statistics

Reporting frequencies for ONJ and AFF ADRs were calculated by the equation:

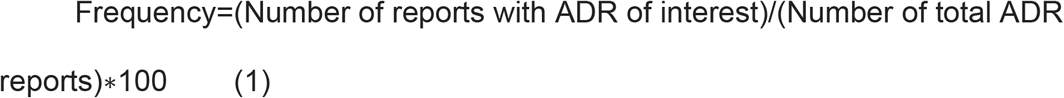

#### Comparative Statistics

The analysis was performed and presented according to established guidelines of pharmacovigilance research established by the FDA and the scientific community. Disproportionality analysis term Reporting Odds Ratio (ROR) was used to differentiate the study from other observational epidemiological studies[49-51].

ADR report rates were compared via the ROR analysis for Figures 1-6 using the following equations:

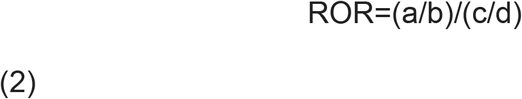

Where:

a. ONJ/AFF cases in exposed group with an AE
b. ONJ/AFF cases in exposed group with no AE
c. ONJ/AFF cases in control group with the AE
d. ONJ/AFF cases in control group with no AE

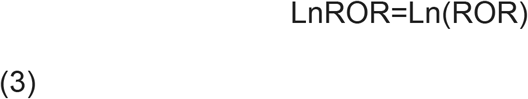

Standard Error of Log Reporting Odds Ratio;

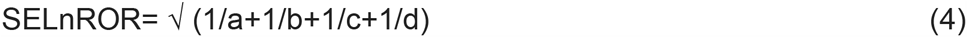

95% Confidence Interval;

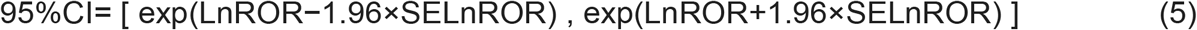

## Results

### Osteonecrosis of the jaw (ONJ)

Patients with osteoporosis-only indication who used bisphosphonates or denosumab had an about hundred-fold higher frequency of ONJ when compared to teriparatide, (reporting odds ratio, ROR, 128.68, 95% confidence interval (CI) [84.45, 196.06]) and (108.74 [71.07, 166.39]) respectively (Fig 2a,2b).

**Fig 2.**
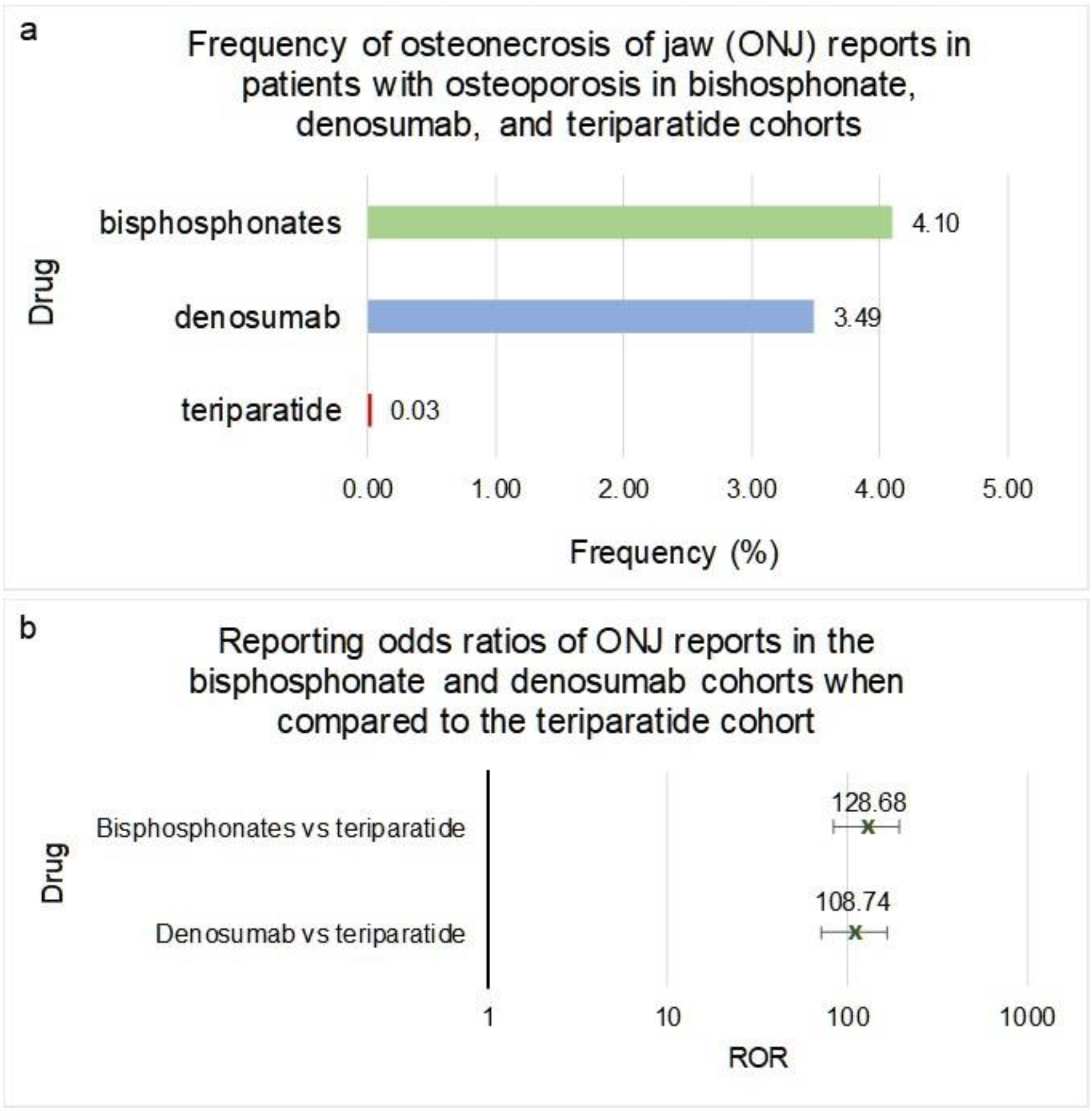
Frequencies and reporting odds ratios (RORs) of osteonecrosis of the jaw (ONJ) adverse events in bisphosphonates, denosumab and teriparatide cohorts. (a) Frequencies of ONJ for patients in FAERS who took bisphosphonates as a class (n=36,527), denosumab (n=18,336) or teriparatide (n=66,173). (b) Reporting odds ratios were calculated comparing adverse event frequencies of bisphosphonates, denosumab and teriparatide patients. Ranges represent 95% confidence intervals (95% CI) (see Methods). X-axis is presented in log scale. Abbreviations: ONJ – osteonecrosis of jaw, ROR – reporting odds ratio.

While in teriparatide comparison the 95%CI ROR ranges of denosumab and bisphosphonate-class overlapped, bisphosphonates as a class, when compared directly with denosumab, had a significantly higher risk of ONJ (1.18 [1.08, 1.30]) (Fig 3a and 3b). However individual bisphosphonate drugs varied in the ONJ RORs vs denosumab. Interestingly, the alendronate cohort had a two-fold increase in risk of ONJ when compared to denosumab (2.00 [1.84, 2.26]) (Fig 3a,3b). On the other hand, ibandronate and zoledronic acid cohorts had significantly lower frequencies of ONJ when compared to denosumab, (0.52 [0.43,0.65]) and (0.61 [0.53, 0.70]) respectively (Fig 3a,3b). The difference in the frequency of ONJ in patients with osteoporosis taking risedronate did not meet the significance criteria when compared to denosumab (1.06 [0.84,1.33]) (Fig 3a,3b).

**Fig 3.**
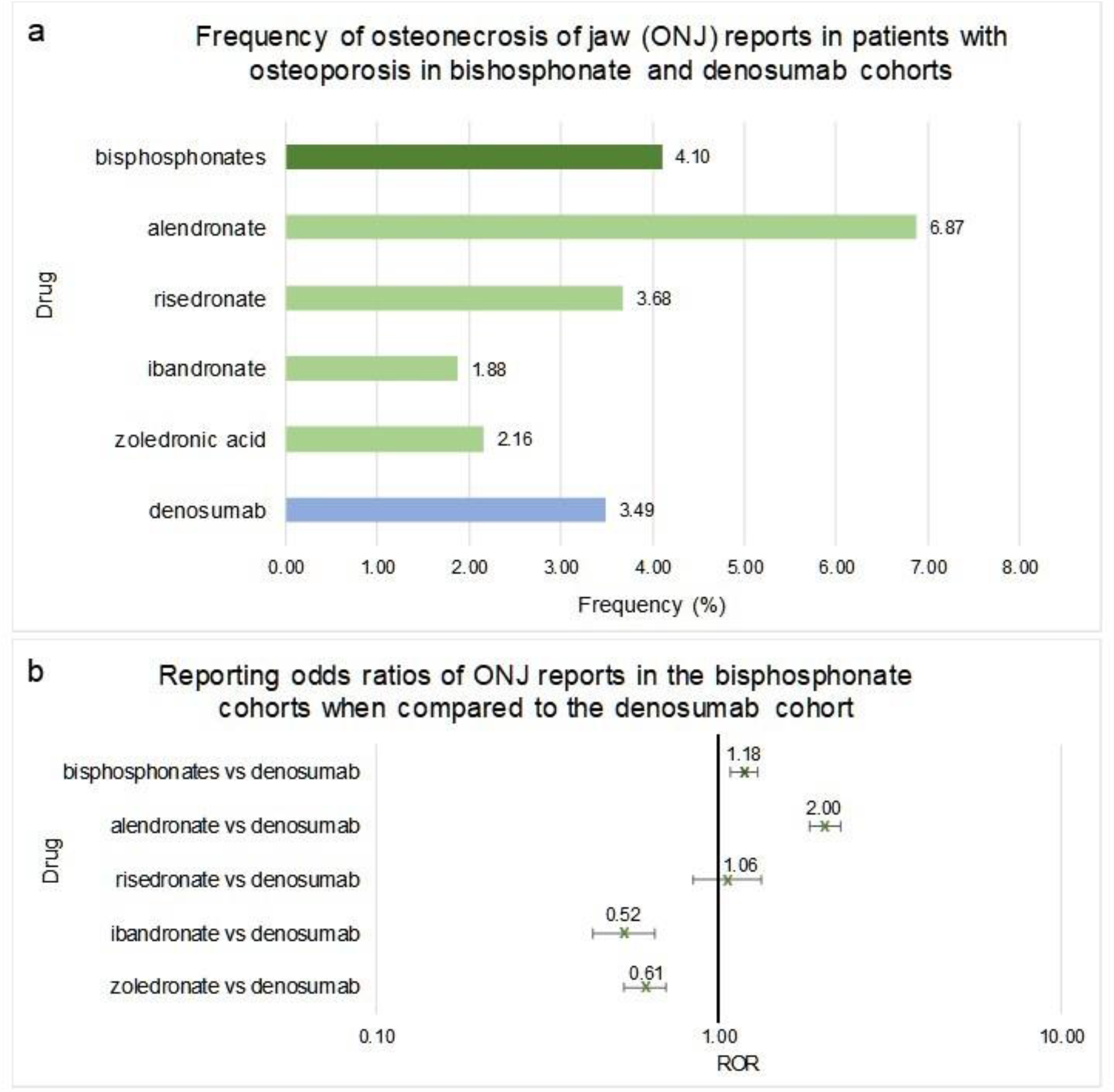
Frequencies and reporting odds ratios (RORs) of osteonecrosis of the jaw (ONJ) adverse events in patients with osteoporosis taking bisphosphonates or denosumab. (a) Frequencies of ONJ for patients in FAERS who took bisphosphonates as a class (n=36,527), individual bisphosphonates: alendronate (n=14,682), ibandronate (n=6,065), risedronate (n=2,309), zoledronic acid (n=13,471), or denosumab (n=18,336). (b) Reporting odds ratios were calculated comparing adverse event frequencies of bisphosphonates and denosumab patients. Ranges represent 95% confidence intervals (95% CI) (see Methods). X-axis is presented in log scale. Abbreviations: ONJ – osteonecrosis of jaw, ROR – reporting odds ratio.

Females who used bisphosphonates as a class or alendronate, as an individual bisphosphonate, had a significant increase in the relative reporting odds ratio of ONJ when compared to denosumab (1.20 [1.10,1.33]) and (2.10 [1.84, 2.30 respectively (Fig 4a,4b). The ibandronate and zoledronic acid cohorts had significantly lower risk of ONJ compared to denosumab (0.55 [0.44, 0.67]) and (0.66 [0.57, 0.76]) respectively (Fig 4a,4b).

**Fig 4.**
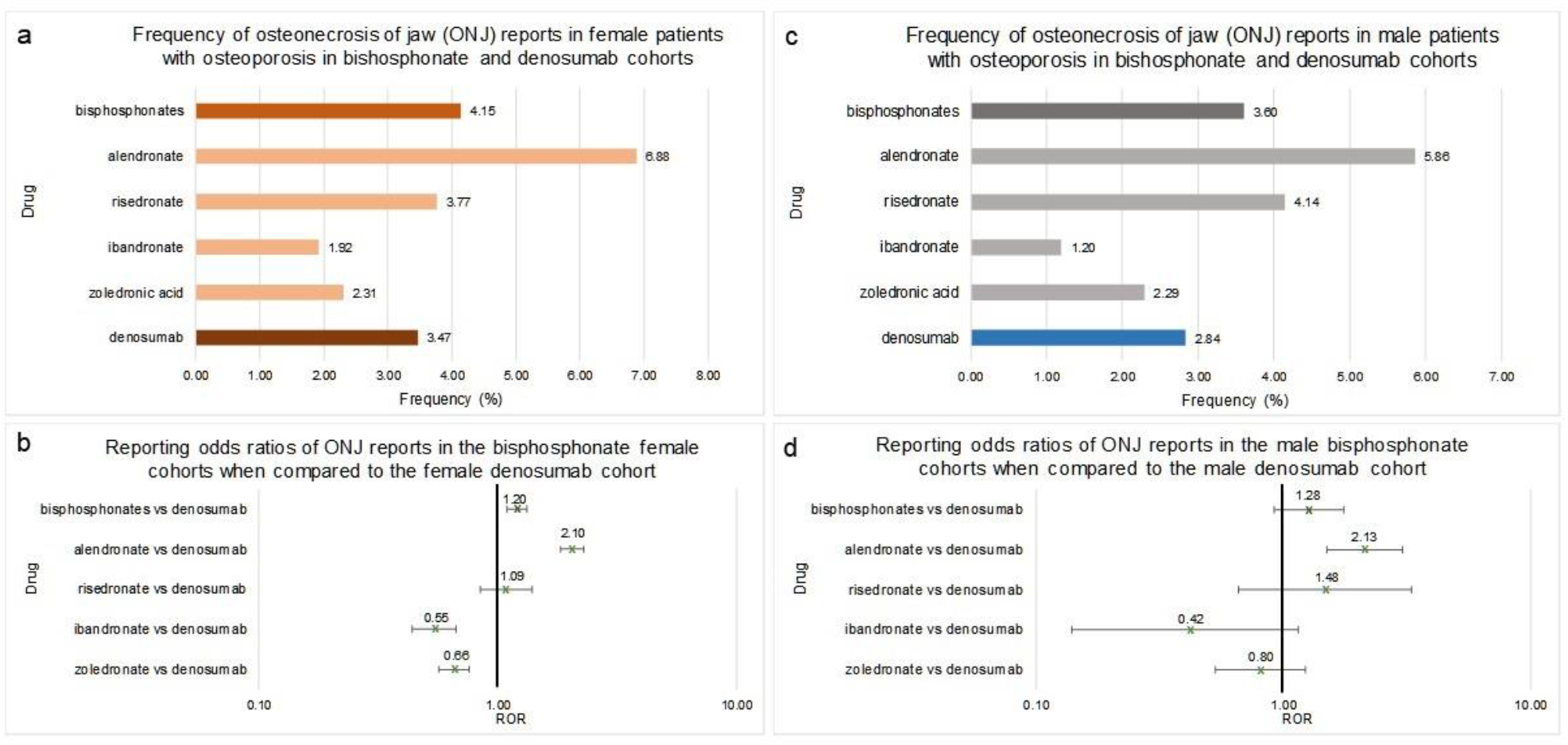
Frequencies and reporting odds ratios (RORs) of osteonecrosis of the jaw (ONJ) in female and male patients with osteoporosis taking bisphosphonates or denosumab. (a) frequencies of ONJ for female patients in FAERS who took bisphosphonates as a class (n=31,834), alendronate (n=12,614), ibandronate (n=5,614), risedronate (n=2,016), zoledronic acid (n=11,590) or denosumab (n=15,690). (b) Reporting odds ratios were calculated comparing adverse event frequencies females taking bisphosphonates or denosumab. (c) frequencies of ONJ for male patients in FAERS who took bisphosphonates as a class (n=3,303), alendronate (n=1,229), ibandronate (n=334), risedronate (n=169), zoledronic acid (n=1,571) or denosumab (n=2,009). (d) Reporting odds ratios were calculated comparing adverse event frequencies females taking bisphosphonates or denosumab. Ranges represent 95% confidence intervals (95% CI) (see Methods). X-axis is presented in log scale. Abbreviations: ONJ – osteonecrosis of jaw, ROR – reporting odds ratio.

Males who used bisphosphonates as a class, ibandronate, risedronate or zoledronic acid had no significant difference in the reporting frequency of ONJ when compared to denosumab (1.28 [0.92, 1.76]), (0.42 [0.14, 1.15]), (1.48 [0.66, 3.30]) and (0.80 [0.53, 1.23]) respectively (Fig 4c, 4d). Similar to females, alendronate male cohort had a significantly higher occurrence and related odds ratio of ONJ when compared to denosumab (2.13 [1.50, 3.04]) (Fig 4c, 4d).

### Atypical femur fracture (AFF)

Patients who used bisphosphonates or denosumab had a significantly higher frequency of AFF when compared to teriparatide. The reporting odds ratio, ROR, for bisphosphonates was 97.16, with 95% confidence interval, CI, [43.32, 217.92]), and for denosumab it was 51.97 [22.71, 118.91] (Fig 5a,5b).

**Fig 5.**
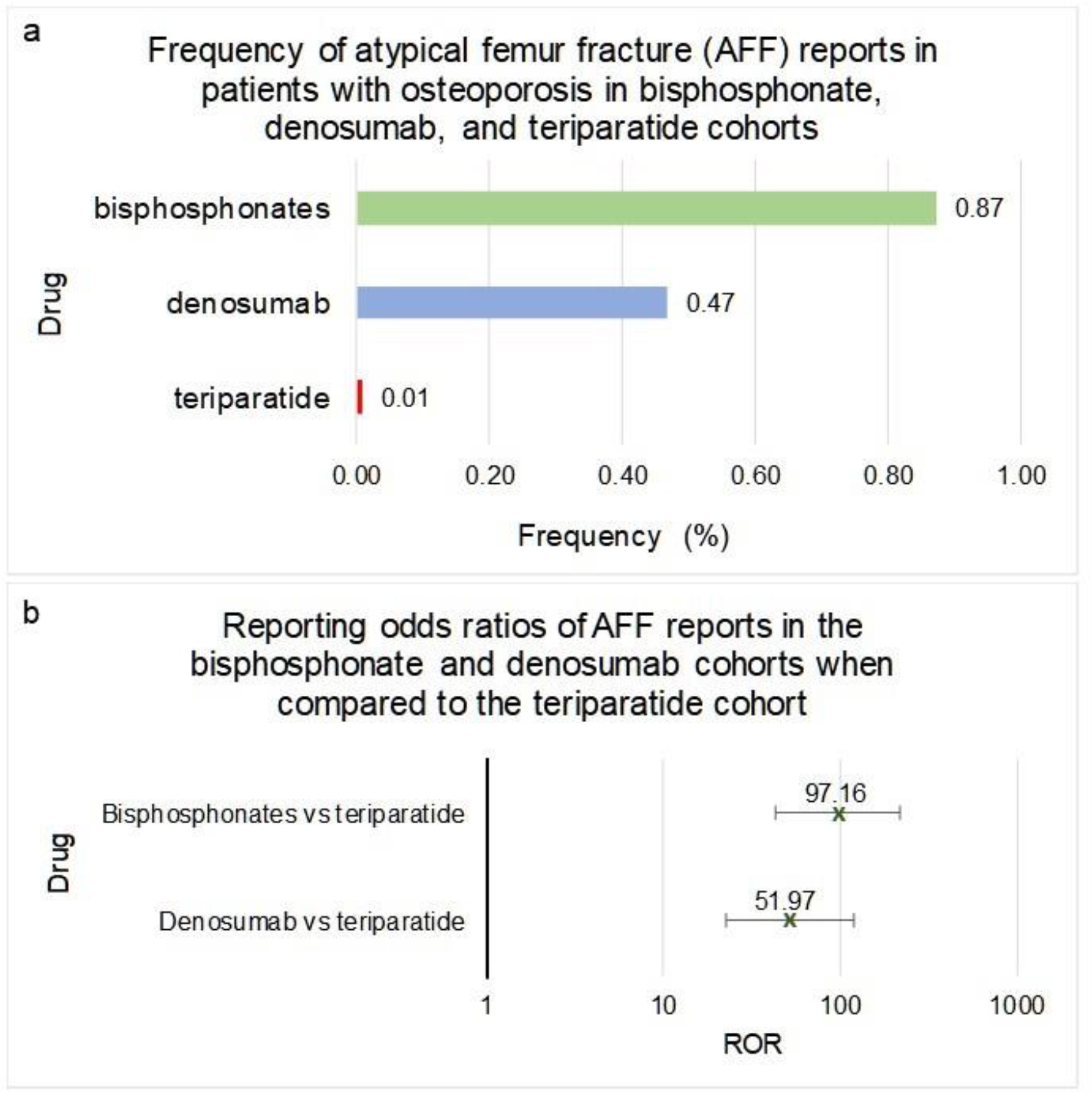
Frequencies and reporting odds ratios (RORs) of atypical femur fracture (AFF) adverse events in bisphosphonates, denosumab and teriparatide cohorts. (a) Frequencies of ONJ for patients in FAERS who took bisphosphonates as a class (n=36,527), denosumab (n=18,336) or teriparatide (n=66,173). (b) Reporting odds ratios were calculated comparing adverse event frequencies of bisphosphonates, denosumab and teriparatide patients. Ranges represent 95% confidence intervals (95% CI) (see Methods). X-axis is presented in log scale. Abbreviations: AFF – atypical femur fracture, ROR – reporting odds ratio.

When compared to denosumab, patients taking bisphosphonates had a significant increase in the frequency of AFF (1.87 [1.47, 2.37]) (Fig 6a,6b). However, the adverse effects between the four bisphosphonates under study differed significantly. The risedronate and alendronate cohorts had significantly higher ROR values of AFF than that of denosumab (6.63 [4.82, 9.12]) and (3.03 [2.36, 3.91]) respectively (Fig 6a,6b). The zoledronic acid cohort had a significantly lower AFF RORs when compared to denosumab, (0.29 [0.18, 0.49]) (Fig 6a,6b). There was no significant difference in the reported frequencies of AFF in patients taking ibandronate when compared to denosumab (ROR = 0.80 [0.51,1.28]) (Fig 6a,6b).

**Fig 6.**
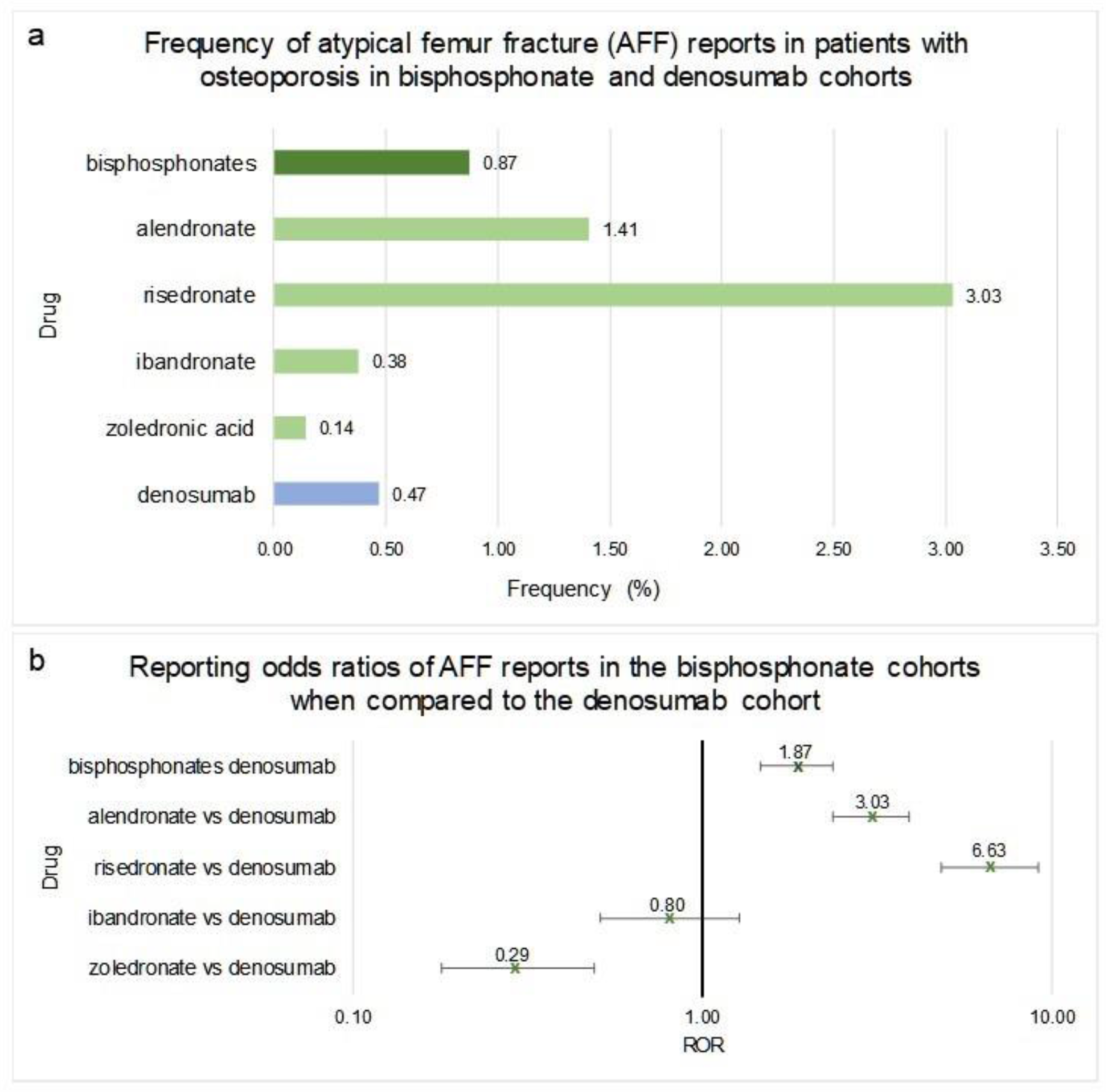
Frequencies and reporting odds ratios (RORs) of atypical femur fracture (AFF) adverse events in patients with osteoporosis taking bisphosphonates or denosumab. (a) Frequencies of ONJ for patients in FAERS who took bisphosphonates as a class (n=36,527), individual bisphosphonates: alendronate (n=14,682), ibandronate (n=6,065), risedronate (n=2,309), zoledronic acid (n=13,471), or denosumab (n=18,336). (b) Reporting odds ratios were calculated comparing adverse event frequencies of bisphosphonates and denosumab patients. Ranges represent 95% confidence intervals (95% CI) (see Methods). X-axis is presented in log scale. Abbreviations: AFF – atypical femur fracture, ROR – reporting odds ratio.

Females who used bisphosphonates as a class, alendronate, or risedronate had a significant increase in the frequency of AFF when compared to denosumab (1.83 [1.47,2.44]), (3.11 [2.38, 4.10]) and (6.58 [4.70, 9.21]) respectively (Fig 7a,7b). The zoledronic acid cohort had a significantly lower frequency of AFF when compared to denosumab (ROR=0.28 [0.17, 0.49]) (Fig 7a, 7b). The ROR value of AFF in female ibandronate patients did not meet the significance criteria when compared to denosumab since the range covered the value of 1 (0.84 [0.53,1.35]) (Fig 7a, 7b). Males who used bisphosphonates as a class, alendronate, or zoledronic acid had no significant difference in the AFF RORs when compared to denosumab (1.13 [0.48, 2.68]), (1.43 [0.52, 3.96]), and (0.47 [0.13, 1.81]) respectively (Fig 7c, 7d). The risedronate cohort had a significantly higher AFF ROR when compared to denosumab (6.63 [4.82, 9.12]) (Fig 7c, 7d). There were no reports of AFF in male patients taking ibandronate for osteoporosis.

**Fig 7.**
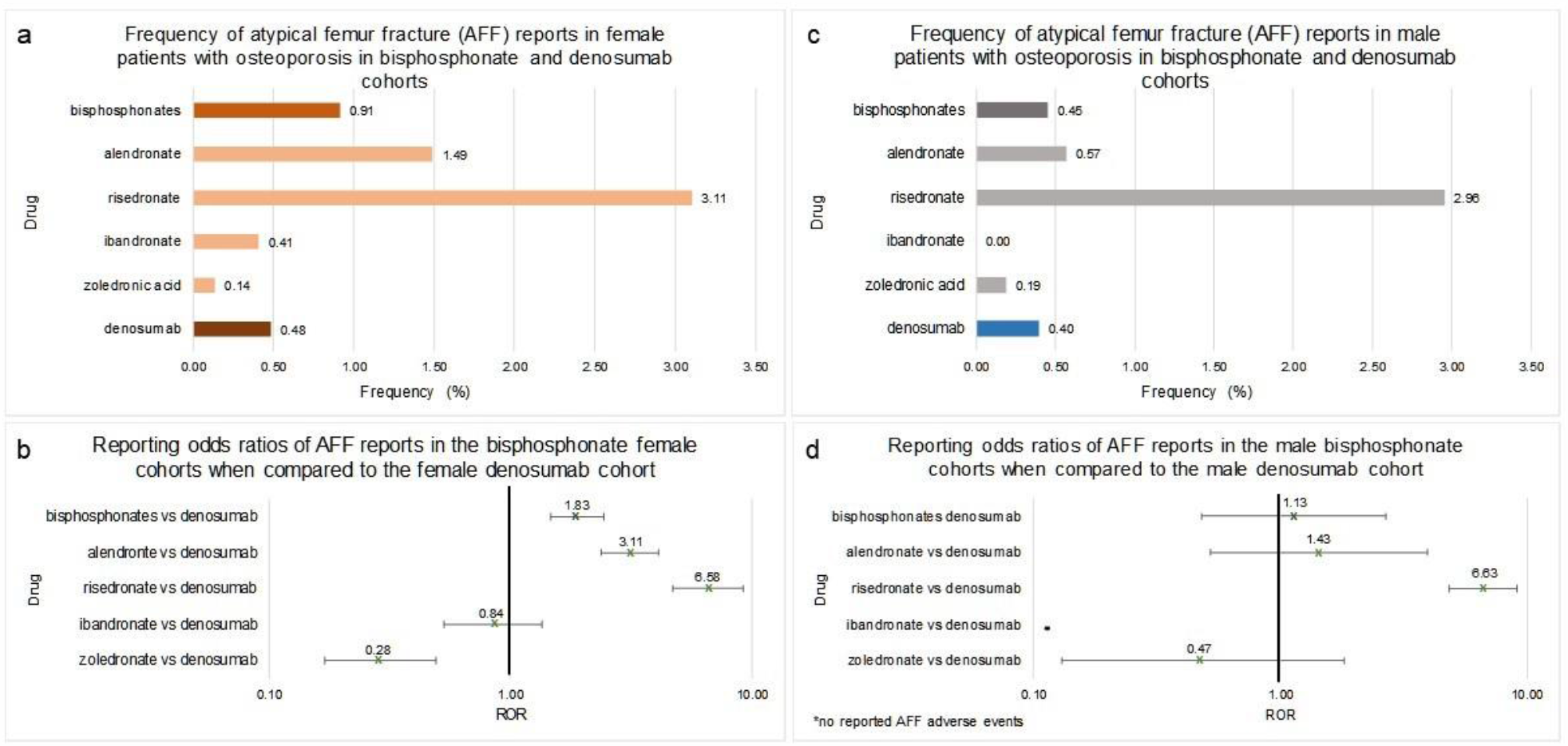
Frequencies and reporting odds ratios (RORs) of atypical femur fracture (AFF) in female and male patients with osteoporosis taking bisphosphonates or denosumab. (a) frequencies of AFF for female patients in FAERS who took bisphosphonates as a class (n=31,879), alendronate (n=12,614), ibandronate (n=5,614), risedronate (2,016), zoledronic acid (n=11,590) or denosumab (n=15,690). (b) were calculated comparing adverse event frequencies females taking bisphosphonates or denosumab. (c) frequencies of AFF for male patients in FAERS who took bisphosphonates as a class (n=3,303), alendronate (n=1,229), ibandronate (n=334), risedronate (n=169), zoledronic acid (n=1,571) or denosumab (n=2,009). (d) RORs were calculated comparing adverse event frequencies females taking bisphosphonates or denosumab. Ranges represent 95% confidence intervals (95% CI) (see Methods). X-axis is presented in log scale. Abbreviations: AFF – atypical femur fracture, ROR – reporting odds ratio.

In a separate analysis we investigated the most common AEs reported alongside AFF and ONJ and observed that: 1) AFF and ONJ co-occurrence was extremely rare and was reported only in 3 out of 18,336 denosumab reports (Supplement Table S1); 2) ONJ co-occurred primarily with dental health and procedure complications, and; 3) AFF most often co-occurred with falls and bone health related AEs (Supplement Table S2).

## Discussion

In this study we quantified the association between individual bisphosphonate use and AFF and ONJ adverse events. We also confirmed the association between bisphosphonates and denosumab exposure and the increased risk of these effects.

To our knowledge this is the first study that used over 12 million reports including 230 thousand osteoporosis reports from the FDA FAERS data (between 2004 and 2019) to evaluate and compare the frequency of rare adverse events associated with the use of three common osteoporosis drug types for the treatment and prevention of osteoporosis.

The risk of ONJ was significantly higher with for both bisphosphonates as a class, and denosumab when compared to teriparatide. However, the adverse effects risk related to each of individual bisphosphonate varied dramatically. Alendronate had the highest frequency of ONJ, and ibandronate had the lowest frequency. Our findings were consistent with those from the study conducted by Zhang and colleagues[52] where they used FAERS reports from the first quarter of 2010 to the first quarter of 2014 and assessed only ibandronate and risedronate. In our study we utilized a much broader data set (2004-2019) and performed a direct pairwise comparison between alendronate, risedronate, ibandronate, zoledronic acid, and denosumab. When analyzed separately, male and female reports yielded similar results with alendronate exhibiting a nearly two-fold increase in risk of AFF.

Osteoporosis patients using bisphosphonates as a class, had higher frequency of AFF when compared to denosumab, and around a hundred-fold higher value when compared to teriparatide. However, similar to the ONJ adverse effect, this difference was not preserved when individual bisphosphonates were analyzed. Patients taking risedronate and alendronate had the highest risk of AFF, while zoledronic acid had the lowest. Our study confirmed the findings of Edwards and colleagues and other studies that showed AFF risk with bisphosphonates[53-55]. There was a significantly higher risk of AFF in females taking alendronate and risedronate, while in male patients only risedronate exhibited significant risk.

Interestingly, we observed a tentative inverse relationship of ONJ and AFF risk with frequency of administration of bisphosphonates: zoledronic acid (once a year), ibandronate (once every three months [IV], or once a month [oral]), risedronate (once a week to once a month), alendronate (once a week)[10-13]. For both side effects the trend in reporting odds ratios was similar for females and males, although in males the association was not always significant due to smaller numbers of reports.

## Conclusion

In our study we observed various levels of risk of ONJ and AFF adverse effects in FAERS reports of individual bisphosphonates with respect to denosumab reports. It may be beneficial to choose zoledronic acid treatment for female patients who are at risk of AFF. In patients at risk of developing ONJ, zoledronic acid may also be the safer option. However, this consideration was restricted to only two adverse effects, further research and controlled trials are needed, and health care providers should use their professional judgment and weigh all the risks and benefits to determine the best treatment option for each specific patient.

### Study limitations

The FAERS reporting system is voluntary indicating that the number of reports available do not represent the number of actual cases and that adverse drug reaction frequencies do not represent actual population incidences. A study done by Alatawi and colleagues using FAERS to found a significant underreporting of adverse events[56]. Additionally, overreporting due to newsworthiness and legal reasons may add noise to the analysis[57, 58]. Absence of comprehensive medical records and demographic variables further limits the extent of our analysis. By using the indication section in the data set, potential comorbidities were excluded, however, due to incomplete reporting, some comorbidities and concurrent medication records may be missing. Although only monotherapy reports were selected for the analysis, some concurrent and over-the-counter medications might have been underreported. That could potentially have introduced error in frequencies and reported odds ratios calculations. As with any association study, causation cannot be established based on RORs. However, analysis of over 230,000 reports provides large scale evidence for rare side effects that may go unnoticed in clinical trials or can be difficult to quantify in smaller observational studies. Despite the limitations, FAERS/AERS remains to be a unique source of population scale data used to identify and quantify rare AEs that can affect patient safety [59].

## Data Availability

Data were obtained from the FDA Adverse Event Reporting System and can be accessed publicly.
The authors confirm that they did not have any special access privileges to these data.

https://www.fda.gov/Drugs/GuidanceComplianceRegulatoryInformation/Surveillance/AdverseDrugEffects/ucm082193.htm

## Declarations

### Ethics Approval

The study used publicly available de-identified data. Institutional Review Board requirements do not apply.

### Availability of data and material

Data were obtained from the FDA Adverse Event Reporting System and can be accessed at https://www.fda.gov/Drugs/GuidanceComplianceRegulatoryInformation/Surveillance/AdverseDrugEffects/ucm082193.htm

The authors confirm that they did not have any special access privileges to these data.

### Code availability

Standard Unix code was used for $ separated .txt file analysis

### Authors’ contributions

T.M. and L.S.A performed the experiments. R.A. and T.M. designed the study and, R.A., L.S.A. and T.M. drafted the manuscript and reviewed the final version. R.A. processed the data set.

## Acknowledgements

We thank Da Shi for contributions to processing the FAERS/AERS data files, demographic analysis, and supporting the computer environment.

